# Laboratory Study of Physical Barrier Efficiency for Worker Protection against SARS-CoV-2 while Standing or Sitting

**DOI:** 10.1101/2021.07.26.21261146

**Authors:** Jacob Bartels, Cheryl Fairfield Estill, I-Chen Chen, Dylan Neu

## Abstract

Transparent barriers were installed as a response to the SARS-COV-2 pandemic in many customer-facing industries. Transparent barriers are an engineering control that are utilized to intercept air traveling between customers to workers. Information on the effectiveness of these barriers against aerosols is limited. In this study, a cough simulator was used to represent a cough from a customer. Two optical particle counters were used (one on each side of the barrier, labeled reference and worker) to determine the number of particles that migrated around a transparent barrier. Nine barrier sizes and a no barrier configuration were tested with six replicates each. Tests of these 10 configurations were conducted for both sitting and standing scenarios to represent configurations common to nail salons and grocery stores, respectively. Barrier efficiency was calculated using a ratio of the particle count results (reference/worker). Barriers had better efficiency when they were 9 to 39 cm (3.5 to 15.5”) above cough height and at least 91 cm (36”) wide, 92% and 93% respectively. Barriers that were 91 cm (36”) above table height for both scenarios blocked 71% or more of the particles between 0.35–0.725 µm and 68% for particles between 1 to 3 µm. A barrier that blocked an initial cough was effective at reducing particle counts. While the width of barriers was not as significant as height in determining barrier efficiency it was important that a barrier be placed where interactions between customers and workers are most frequent.

## Introduction

SARS-CoV-2, a novel virus that causes COVID-19, is transmitted mostly through close contact. Close contact is defined by the CDC as: “someone within six feet of an infected individual for a cumulative total of 15 minutes or more within a 24 hour period (CDC 2021a)”. The cumulative time of exposure to infected individuals has been associated with COVID-19 risk (CDC 2020b; Pringle et al. 2020). Employees in the service industries that spend extended (e.g., manicures take 30 to 60 minutes) time with multiple customers, such as nail technicians and cashiers, are at greater risk than other workers (CDC 2020d; Faces Spa 2020). Despite the availability of safe and effective vaccines against COVID-19, employee exposure risk remains a concern because COVID-19 cases continue to occur (including the now predominant Delta variant), not everyone has been vaccinated, and mask mandates have been reduced (CDC 2021b; Markowitz 2021).

Transparent barriers, generally made of polycarbonate or polymethylmethacrylate (PMMA), are recommended by the Centers for Disease Control and Prevention (CDC) (CDC 2020a) and the Occupational Safety and Health Administration (OSHA) (OSHA 2021) as engineering controls to limit the transmission of SARS-CoV-2 to workers. Evidence-based recommendations are needed on the effectiveness of barriers and size requirements for employees in customer-facing industries such as the 155,300 nail salon technicians, 3.6 million grocery and convenience store cashiers, and other workers (Bureau of Labor Statistics 2021). The purpose of barriers is to reduce the number of inhalable (<10 µm aerodynamic equivalent diameter (AED)) and respirable (<4 µm AED) particles that could migrate from an infected customer to a worker on the opposite side of a barrier, or from an infected worker to a customer (World Health Organization 2020). How barriers affect particle migration at the submicron fraction is vital as research indicates that infectious virions can be contained in particles smaller than 1 µm (Santarpia et al. 2020).

Transparent barriers provide desirable qualities as an intervention method. These barriers serve as a semi-permanent engineering control, enforce physical distancing, are easily sanitized, and protect from other aerosol-transmitted diseases (Environmental Health and Safety University of Washington 2020). However, evidence is limited on the effectiveness of transparent barriers for preventing SARS-CoV-2 transmission and the appropriate size of barriers to provide the best efficiency for particle reduction (CDC 2020c; Lindsley et al. 2013). For instance, the University of Washington proposed barrier heights, from the floor to the top of the barrier, based on the tallest individuals (Environmental Health and Safety University of Washington 2020).

The objective of this laboratory study was to determine the efficiency of transparent barriers in blocking airborne particles and to evaluate efficiency against barrier size. Most particles that were produced by this study’s cough simulator were in the respirable range (<4 µm) which travel into the lungs during breathing; some particles were in the inhalable range (<10 µm) which likely deposit into the nose, mouth, and throat. The size ranges of particles generated by the simulator in this study are similar to that of an adult human cough, with a count median diameter between 0.57 and 0.71 μm (average 0.63 μm, SD 0.05) (Lindsley et al. 2013). Size ranges for individual coughs vary greatly and have been measured from 0.01 to 500 µm (Gralton et al. 2011). Although larger particles (up to 100 µm) can be inhaled, most are not. Cloth masks, if worn properly, reduce the number of larger particles (>3 µm) both released and inhaled by the wearer, but for smaller particles additional protection is needed (Akhtar et al. 2020; Lindsley et al. 2021).

## Materials and Methods

### Cough Simulator

The cough simulator is made from a metal bellows and automated by a linear motor (Model STA2506; Copley Controls, Canton, MA). It has been described previously (Lindsley et al. 2021; Lindsley et al. 2019; Lindsley et al. 2014; Lindsley et al. 2013). Briefly, the cough aerosol expelled by the simulator is produced from a 28% Potassium Chloride (KCl) solution generated in a single-jet Collison nebulizer (ARGCNB1, CH Technologies (USA), Inc. Westwood, NJ) under 172 Kilopascal (25 pounds/in2). The expelled aerosol for each “cough” had a peak flow rate of 11 L/s and a volume of 4.2 L. The flow rate and volume were similar to influenza patients as determined by Lindsley et al. (Lindsley et al. 2012).

### Transparent Barriers

The transparent barriers were clear acrylic sheets (OPTIX, Plaskolite, Columbus, OH). The acrylic sheets were 0.3 cm (0.125”) thick and cut into nine sizes. The heights were 61, 91 or 122 cm (24, 36, and 48”), widths were 61, 91 or 122 cm (24, 36, and 48”) for nine test sizes. A rectangular opening, 10 cm (4”) high by 31 cm (12”) wide, was cut into the bottom center of each test barrier to represent access for transactions. A control trial was conducted without the use of a barrier for use as a comparison for calculating efficiency. For data analysis, the vertical height between the top of the barrier and the mouth of the cough manikin was used.

### Aerosol Counts

The aerosols produced by the cough simulator were counted by factory calibrated GRIMM optical particle counters (OPC, Model 1.108; Grimm Technologies, Inc., Douglasville, GA). Two OPCs were used for each experimental run, denoted as reference OPC and worker OPC. Prior to sampling, each OPC was synced to the same computer to ensure matching times for each data point collected. Prior to initial data collection both OPCs displayed similar pre-and post-cough particle count measurements. The reference OPC was placed 30.5 cm (12”) from the front of the “mouth” of the cough simulator on the same side of the barrier with the inlet port directly in line at the same height. The worker OPC was placed on the far side of the barrier. The OPCs collected data every six seconds for particles in the size range from 0.35 to 22.0 µm in 15 size bins. For analysis, particle sizes were categorized into small (0.35–0.725 µm), moderate (0.9–2.5 µm), large (3.5–6.25 µm) and extra-large (8.75–22.5 µm).

### Isolation room

The experiment was performed in an isolation room (Figure 1). This isolation room was originally designed as a model hospital airborne infection isolation room (Thatiparti, Ghia and Mead 2016). Physical measurements of the isolation room were 4.9 m (16’) by 4.3 m (14.1’) with a ceiling height of 2.4 m (8’) for a total of 58 m^3^ (2057 ft^3^). The isolation room ventilation rate was set to two air changes per hour (ACH) and confirmed by an initial sulfur hexafluoride (SF6) decay test, which indicated an ACH of 2.40, and multiple measurements throughout the study using a balometer (TSI/ALNOR EBT731, Shoreview, MN), which ranged from 1.81 to 2.11 ACH. Around 2 ACH is the calculated value for office spaces (ASHRAE 2004). The supply air vents had measured readings, on average, of 17 L/s (36 cfm) for the vent behind the cough manikin and 16 L/s (34 cfm) for the vent opposite the manikin. The return air vents had measured exhaust readings, on average, of 9.9 L/s (21 cfm) for the vent opposite the manikin and 11.3 L/s (24 cfm) for the vent in the supply closet, which was open, as shown in Figure 1. A smoke generator showed that the air generally flowed from behind the cough simulator past the barrier. Using the physical dimensions and airflow measurements, an air change rate (ACH) was calculated by dividing the greater of total exhaust or supply (m^3^/hr) by the area volume (m^3^).

**Figure 1.**
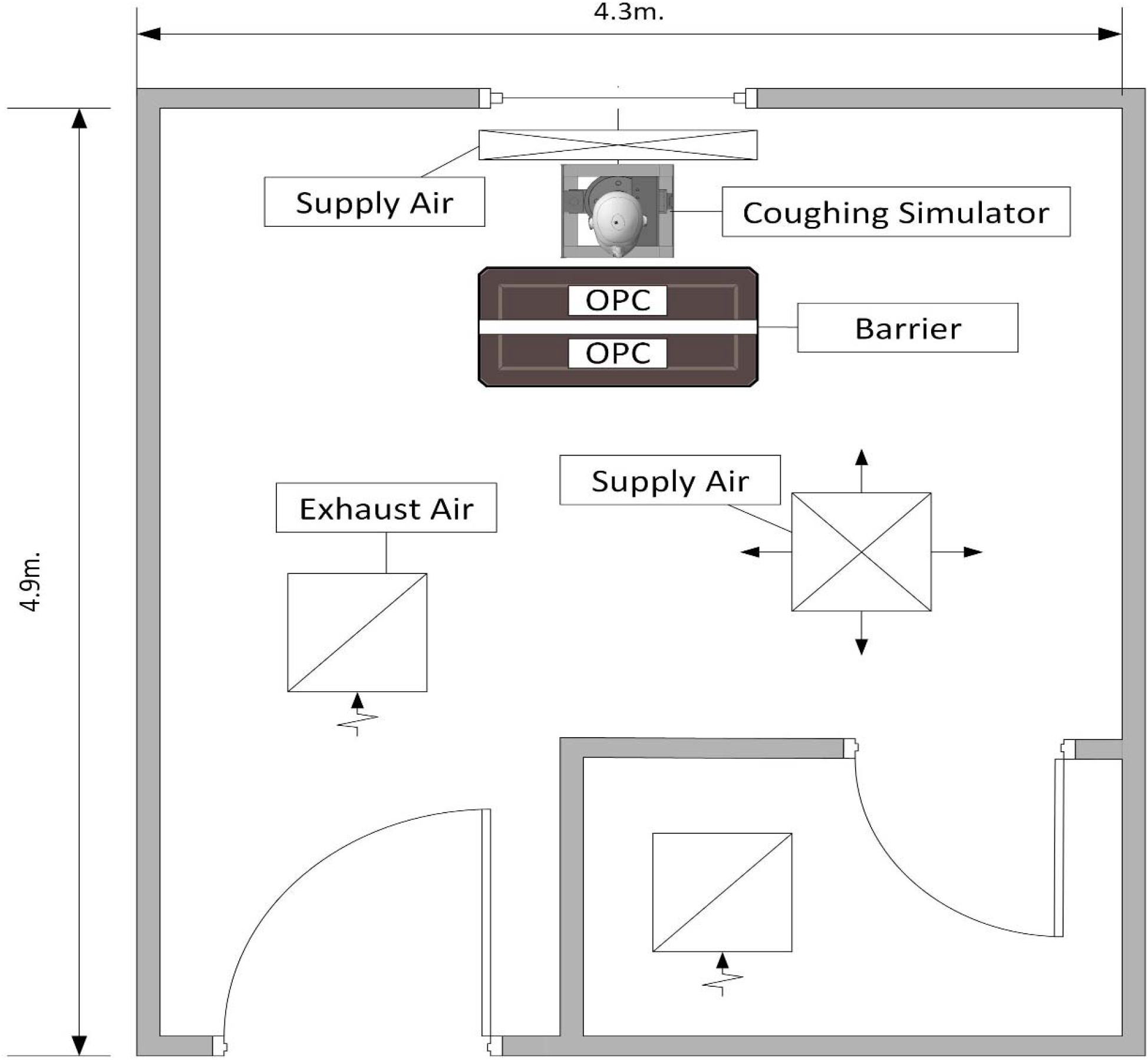
Isolation Room 4.9 m (16’) long, 4.3 m (14 ‘ 2 ”) wide and 2.5 m (8’) tall. Note: the exhaust and supply vents were all on the ceiling (2.5 m). The supply closet door was open during the study.

### Seated and standing workstation design

Two scenarios were analyzed to represent typical interactions: seated and standing (Figures 2 and 3). A seated scenario represented an interaction between a worker and customer seated at a table, such as at a nail salon manicurist table. The standing scenario represented an interaction between a worker and customer standing at a counter, such as at a cashier counter or grocery checkout. The cough simulator was placed 76 cm (30”) from the barrier and 30.5 cm (12”) from the reference OPC. The worker OPC was placed 46 cm (18”) from the barrier on the opposite side.

**Figure 2.**
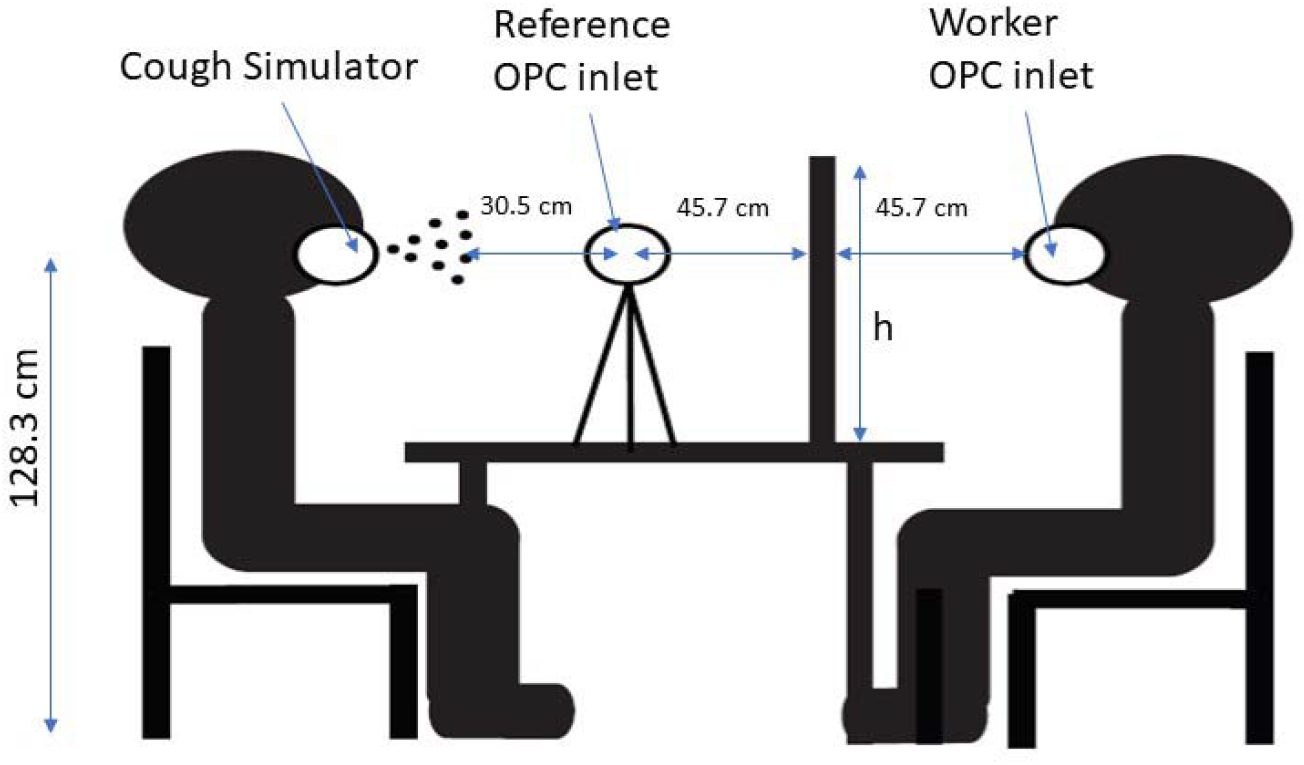
Seated Interaction shown with a cough simulator, worker, and reference optical particle counters and transparent barrier. h indicates the height of the barrier, which was 61, 91 or 122 cm (24, 36 and 48”) from the table. The barrier widths, not shown, were 61, 91 or 122 cm (24, 36, and 48”). Table dimensions were 76 cm (30”) tall, 61 cm (24”) wide, and 102 cm (40”) long. A rectangular opening was cut into the bottom center of each barrier that was 10 cm (4”) tall and 31 cm (12”) wide.

**Figure 3.**
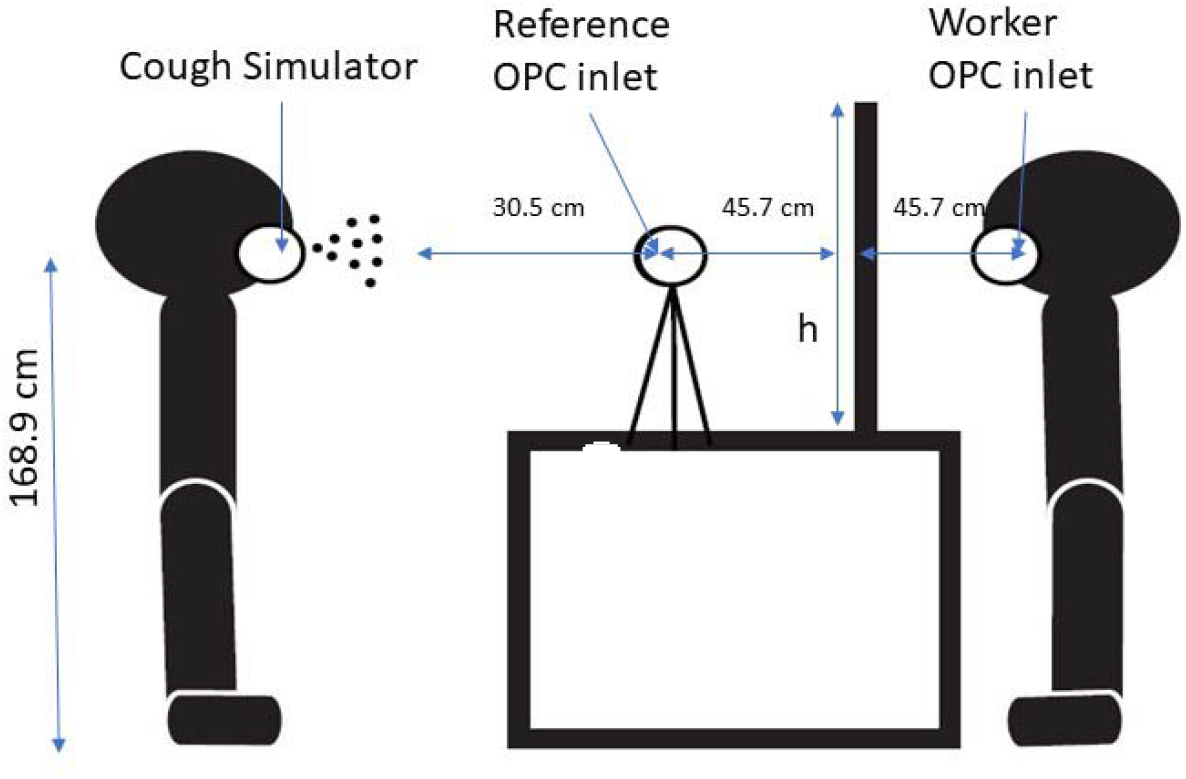
Standing interaction with cough simulator, worker, and reference optical particle counters, and transparent barrier. h indicates the height of the barrier, which is 61, 91 or 122 cm (24, 36, and 48”) from the table. The barrier widths, not shown, were 61, 91 or 122 cm (24, 36, and 48”). Table dimensions were 91 cm (36”) high, 61 cm (24”) wide, and 102 cm (40”) long. A rectangular opening was cut into the bottom center of each barrier that was 10 cm (4”) tall and 31 cm (12”) wide.

It is expected that customers and employees shift positions throughout an interaction. It was not feasible to represent this movement due to variation between individuals and situations. The OPCs were attached to tripods to elevate them to the correct height. The cough simulator was elevated by a constructed 80/20® (Columbia City, IN) aluminum frame.

The sitting scenario represented an exposure at a salon while conducting a manicure. Since more women than men visit salons (72% compared to 52%), a woman’s average height was used for the sitting workstation scenario (Sharma et al. 2018). The cough simulator was positioned at a seated mouth height of a 95^th^ percentile woman. This height represented an increased risk of exposure—a taller person would cough over a shorter barrier. The heights for the worker OPC and the cough simulator were determined from anthropometric data from the American Industrial Hygiene Association (AIHA) Engineering Reference Manual (Estill 1999). Figure 2 shows the heights at which each instrument was placed for the sitting scenario. To calculate cough height, seated eye height (79 cm, 31”) was added to the sitting knee height (55 cm, 21.5”) and 5 cm (2”) were subtracted to estimate the mouth position (128 cm, 50.5”). In the sitting scenario, the table was 76 cm (30”) high, 61 cm (24”) wide, and 102 cm (40”) long, similar to several commercial manicurist tables (Easy Nail Tech). Nail salon workstations are, however, diverse. The vertical distances from the top of the barrier to the mouth of the coughing manikin were 9, 39 and 70 cm (3.5, 15.5, and 27.5”) for the three barrier heights.

For the standing workstations, the cough simulator was placed at a height chosen based on the 95^th^ percentile height for men (Estill 1999). Male stature at the 95^th^ percentile was considered the highest risk of exposure. To estimate mouth position, 15 cm (6”) was subtracted from male stature (184 cm, 72.5”) for a cough height of 169 cm (66.5”). The counter was 91 cm (36”) high with a depth of 61 cm (24”) based on the American National Safety Institute (ANSI) and American with Disabilities Act (ADA) countertop requirements for sales counters (United States Access Board ; US Department of Justice 1999). The vertical distance between the top of the barrier to the mouth of the coughing manikin was 16.5 cm below the mouth, 14 and 44.5 cm above the mouth (−6.5, +5.5, and +17.5“ from mouth height).

### Study Trials

Each trial consisted of three 10-minute phases. During the first phase background particle counts were measured. During the second phase particle counts were obtained while the cough simulator was activated to produce two coughs. The final phase was used to clean the air using a portable high-efficiency particulate air (HEPA) filtered unit (VACOMAGUS, Atrix, Burnsville, MN) at 280 cubic feet per minute (cfm) to reduce aerosols in the room before the next background measure was taken. For each scenario (sitting and standing) the order of barriers was randomized. Six replicates of each of the ten barrier conditions (nine barriers and no barrier) were performed resulting in 60 trials for each scenario.

### Statistical analysis

Summary statistics were provided as median and range for the variable of interest, efficiency, with right-skewed distribution. Efficiency was quantified using the ratios of the reference OPC count to the worker OPC count among ten barrier conditions to determine the percentage reduction. Ratios were adjusted by subtracting the background particle counts.

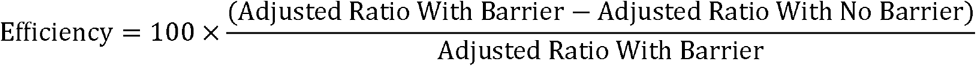

A median regression was carried out to estimate differences in the medians of the outcome or dependent variable, i.e., efficiency, and to determine how two independent variables, i.e., barrier width and height, in combination, influence the outcome in small, moderate, and large-sized particles and in each scenario. Corresponding 95% confidence intervals (CI) and p-values were also provided. Median regression is recommended when the outcome does not follow normal distribution or potential outliers are found to influence the mean relative to the median, both of which were existing for our data (Koenker and Hallock 2001). Additionally, multiple comparisons were performed to investigate significant differences of the outcome variable among all possible combinations of barrier settings. All statistical tests were two-sided at the 0.05 significance level and analyses were conducted in SAS version 9.4 (SAS Institute, Cary, NC).

## Results

Median and range for efficiency with right-skewed distributions are provided (Table 1). For the small sized particles (0.35–0.725 µm), barrier median efficiencies ranged from 71% to 86% for barriers where the top was at least 14 cm (5.5”) above cough height and all widths when compared to no barrier in the standing scenario, while the range was 83% to 93% for barriers at all heights and widths in the sitting scenario. For moderate sized particles (0.9–2.5 µm), barrier median efficiencies ranged from 68% to 87% for barriers at least 14 cm (5.5”) above cough height for standing and from 71% to 94% for barriers at least 9 cm (3.5”) above cough height for sitting. While standing, the 61 cm (24”) high barrier was 16.5 cm (6.5”) below cough height, had low efficiency, and was not significantly different from the no barrier configuration. With respect to large sized particles (3.5–6.25 µm), the median efficiencies for barriers that were above cough height and at least 91 cm (36”) wide were 61% or greater for standing and 69% or greater for sitting (Table S1). Figure 4 presents the geometric means (GMs) of particle counts by category size and OPC location for one randomly selected scenario. The GMs of reference OPC were greater than the values of worker OPC in each scenario. The smallest particles were the most abundant, and differences between reference and worker OPC were greatest for the small and moderate sized particles.

**Table 1.** Collection efficiency by particle size in standing and sitting scenarios.

|  | Small Sized Particles (0.35µm – 0.725µm) |  |  | Moderate Sized Particles (0.9µm – 2.5µm) |  |
| --- | --- | --- | --- | --- | --- |
|  | Number of Experiments | Median Efficiency (Range) | P-value | Median Efficiency (Range) | P-value |
| Barrier in Standing Scenario |  |  |  |  |  |
| No Barrier | 6 | Reference |  | Reference |  |
| Height (H) from Cough |  |  |  |  |  |
| -16.5 cm (-6.5") H | 18 | -1.14 (-551.6 – 77.12) | 0.7869 | -29.46 (-4206 – 79.86) | 0.2612 |
| +14 cm (5.5") H | 18 | 82.00 (41.09 – 96.01) | <.0001 | 74.91 (-324.8 – 94.94) | 0.0007 |
| +44.5 cm (+17.5") H | 18 | 84.46 (10.87 – 96.82) | <.0001 | 76.35 (-430.5 – 96.09) | 0.0008 |
| Width (W) Excluding -16.5 cm (-6.5") H |  |  |  |  |  |
| 61 cm (24") W | 12 | 76.91 (10.87 – 96.01) | <.0001 | 73.02 (-430.5 – 94.80) | <.0001 |
| 91 cm (36") W | 12 | 85.81 (41.09 – 96.82) | <.0001 | 83.09 (-148.0 – 96.09) | <.0001 |
| 122 cm (48") W | 12 | 81.25 (44.68 – 95.20) | <.0001 | 69.54 (-324.8 – 94.94) | 0.0001 |
| Height (H) * Width (W) |  |  |  |  |  |
| -16.5 cm (-6.5") H * 61 cm (24") W | 6 | 5.75 (-185.8 – 72.92) | 0.5369 | -4.87 (-1012 – 79.86) | 0.7870 |
| -16.5 cm (-6.5") H * 91 cm (36") W | 6 | -6.20 (-551.6 – 77.12) | 0.8416 | -10.40 (-4206 – 67.51) | 0.7910 |
| -16.5 cm (-6.5") H * 122 cm (48") W | 6 | -17.87 (-233.4 – 70.22) | 0.7598 | -72.41 (-929.2 – 54.48) | 0.3895 |
| +14 cm (5.5") H * 61 cm (24") W | 6 | 81.34 (59.97 – 96.01) | <.0001 | 76.17 (49.95 – 94.80) | 0.0107 |
| +14 cm (5.5") H * 91 cm (36") W | 6 | 84.25 (41.09 – 94.47) | <.0001 | 81.14 (-148.0 – 92.96) | 0.0148 |
| +14 cm (5.5") H * 122 cm (48") W | 6 | 78.66 (44.68 – 95.20) | <.0001 | 67.90 (-324.8 – 94.94) | 0.0545 |
| +44.5 cm (+17.5") H * 61 cm (24") W | 6 | 70.88 (10.87 – 93.86) | <.0001 | 70.61 (-430.5 – 91.73) | 0.1274 |
| +44.5 cm (+17.5") H * 91 cm (36") W | 6 | 86.32 (80.70 – 96.82) | <.0001 | 86.59 (1.06 – 96.09) | 0.0039 |
| +44.5 cm (+17.5") H * 122 cm (48") W | 6 | 83.02 (61.01 – 94.23) | <.0001 | 72.52 (-228.9 – 93.00) | 0.0224 |
| Barrier in Sitting Scenario |  |  |  |  |  |
| No Barrier | 6 | Reference |  | Reference |  |
| Height (H) |  |  |  |  |  |
| +9 cm (3.5") H | 18 | 87.99 (41.32 – 98.19) | <.0001 | 87.11 (-50.56 – 98.44) | <.0001 |
| +39 cm (15.5") H | 18 | 91.33 (64.88 – 98.76) | <.0001 | 93.23 (13.68 – 99.02) | <.0001 |
| +70 cm (27.5") H | 18 | 89.61 (65.35 – 97.71) | <.0001 | 86.91 (6.87 – 98.45) | <.0001 |
| Width (W) |  |  |  |  |  |
| 61 cm (24") W | 18 | 88.19 (67.30 – 97.08) | <.0001 | 84.97 (-34.82 – 98.04) | <.0001 |
| 91 cm (36") W | 18 | 91.77 (65.35 – 98.76) | <.0001 | 92.95 (7.13 – 98.77) | <.0001 |
| 122 cm (48") W | 18 | 89.30 (41.32 – 98.45) | <.0001 | 86.67 (-50.56 – 99.02) | <.0001 |
| Height (H) * Width (W) |  |  |  |  |  |
| +9 cm (3.5") H * 61 cm (24") W | 6 | 83.18 (73.79 – 92.77) | <.0001 | 70.72 (-34.82 – 89.64) | <.0001 |
| +9 cm (3.5") H * 91 cm (36") W | 6 | 91.96 (85.64 – 98.19) | <.0001 | 94.12 (82.77 – 98.44) | <.0001 |
| +9 cm (3.5") H * 122 cm (48") W | 6 | 86.16 (41.32 – 97.62) | <.0001 | 84.11 (-50.56 – 98.13) | <.0001 |
| +39 cm (15.5") H * 61 cm (24") W | 6 | 90.55 (77.33 – 97.08) | <.0001 | 93.74 (79.39 – 97.97) | <.0001 |
| +39 cm (15.5") H * 91 cm (36") W | 6 | 92.66 (77.97 – 98.76) | <.0001 | 93.15 (42.21 – 98.77) | <.0001 |
| +39 cm (15.5") H * 122 cm (48") W | 6 | 90.65 (64.88 – 98.45) | <.0001 | 90.38 (13.68 – 99.02) | <.0001 |
| +70 cm (27.5") H * 61 cm (24") W | 6 | 88.94 (67.30 – 96.89) | <.0001 | 85.81 (42.42 – 98.04) | <.0001 |
| +70 cm (27.5") H * 91 cm (36") W | 6 | 89.12 (65.35 – 97.71) | <.0001 | 86.91 (7.13 – 97.84) | <.0001 |
| +70 cm (27.5") H * 122 cm (48") W | 6 | 91.33 (69.56 – 97.66) | <.0001 | 89.26 (6.87 – 98.45) | <.0001 |

**Figure 4.**
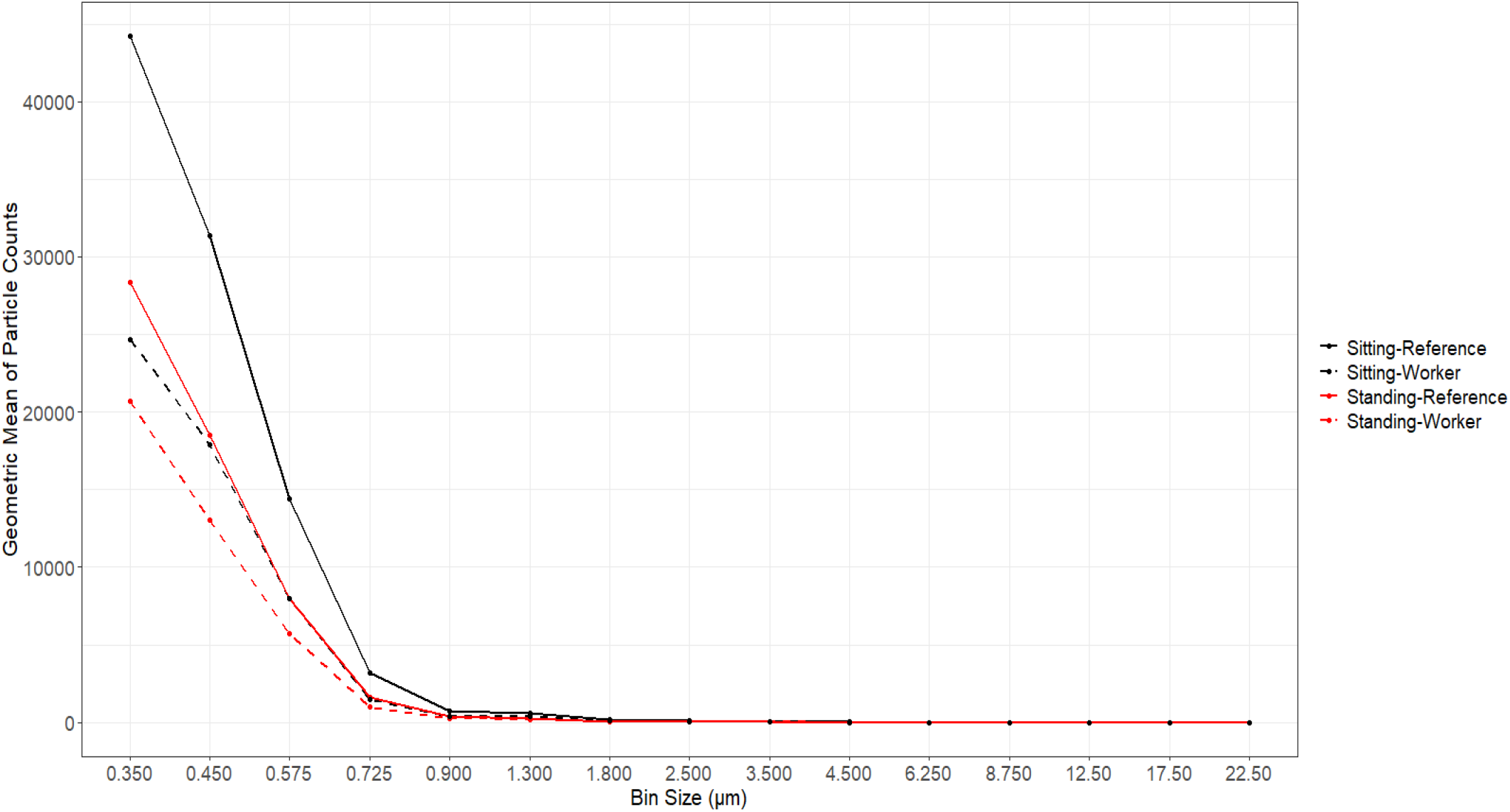
Geometric means of particle counts by bin size and OPC location for the configuration of 14 cm (5.5”) above cough height and 91 cm (36”) width in standing, and 39 cm (15.5”) above cough height and 91 cm (36”) width in sitting.

For the smallest particles (Table S2), the barriers above cough height blocked aerosols by 76 to 90% compared with no barrier (all p-values < 0.0001). For the standing scenario, the efficiencies of all barriers that were at least 14 cm (5.5“) above cough height did not significantly differ from each other. As expected, the barrier that was 16.5 cm (6.5”) below cough height in the standing scenario was not significantly different from no barrier. The results of efficiencies for moderate sized particles are provided in Table S3. Most barriers that were at least 14 cm (5.5“) above cough height in the standing scenario significantly blocked the aerosols relative to no barrier. In the sitting scenario, all barrier settings with moderate size outperformed no barrier by 86 to 93% (p-values < 0.0001).

## Discussion

Physical transparent barriers provide benefits for workers by reducing the number of particles from a customer’s cough or breath that make it to the worker. This laboratory simulation showed that barriers reduced particle count by at least 71% for the smallest particles when located at least 9 cm (3.5”) above cough height.

To be effective a barrier needs to be above cough height so that it can block the exhaled air from a cough. Our analysis showed that the best performing barriers were: (1) at least 9 cm (3.5 “) above cough height, (2) preferably 15 cm (5.5”) above cough height while standing and (3) 39 cm (15.5”) above cough height while sitting. Barriers that were shorter than cough height did not protect and were not different than not using a barrier. The tallest barriers 122 cm (48”) above table height that were used in both scenarios did not provide improved efficiency compared to the 91 cm (36”) tall barriers; there was no added benefit from increasing the barrier height beyond 91 cm (36”).

When considering barrier width, we excluded barriers that were below cough height for the standing scenario. The 91 cm (36”) wide barriers performed better than the 61 cm (24”) wide barriers for small and moderate sized particles in both the sitting and standing scenarios. The 91 cm (36”) wide barrier performed statistically better than the 122 cm (48”) wide barriers for moderate sized particles. There was not a statistically significant difference between barrier widths of 91 cm (36”) and 122 cm (48”) for the small sized particles. Although all the barrier widths were more efficient than no barrier, 91 cm (36”) wide barriers were preferred because they were 3 to 13% more efficient than the 61 cm (24”) and 122 cm (48”).

It has been shown that airflow is impacted by vent and barrier placement (Lee and Awbi 2004). Ventilation configurations will vary greatly at workplaces. Larger barriers were not shown to be statistically better than smaller barrier sizes in our study. For both height and width, the middle-sized barriers had greater efficiencies and sometimes efficiencies that were statistically greater than the largest heights and widths. The largest barriers may have interfered with the room air movement. In this study, air was moving in the room at approximately two ACH, typical for a store or office (ASHRAE 2004). The largest barrier may have reduced mixing and may have entrained the exhaled cough to move in the direction that was the least blocked. For instance, a wider barrier may have forced the exhaled cough to go over a short height. Proper ventilation will aid in preventing smaller particles, which remain in the air longer, from remaining in one location.

To provide worker protection, it is preferred that barriers encompass the location where a customer primarily interacts with a worker. For example, if a customer at a grocery store checkout needs to load groceries and then make a payment, it is preferred that barriers are erected at each interaction location. A good measure to ensure the greatest range of protection for height is to use the 95^th^ percentile of height for males for both sitting and standing interactions. The 95^th^ percentile cough height for U.S. males is 168.9 cm (66.5“). Therefore, the top of the transparent barriers for standing interactions shown to be the most effective in this study was 183 cm (72“) from the floor. For sitting interactions at nail salons, the barrier height shown to be the most effective in this study was 140 cm (55”) from the floor based on the 95^th^ percentile for sitting height for females with a chair height of 56 cm (22”) (Estill 1999). Female height was used because of the greater percentage of female workers and customers in the nail salon industry (Sharma et al. 2018). However, to offer the optimal community protection, the sitting barrier heights of 150 cm (59”) from the floor are preferred as they account for the 95^th^ percentile sitting height for males using a 58 cm (23”) chair height.

There are some limitations in this experiment. First, all experiments were performed in an isolation room that provided consistent ventilation rates for comparison which may not be representative of occupational settings. Further studies with other general air movement patterns are needed. There is a wide variation in workstation configurations. The scenarios used here do not accurately reflect all real-world situations and a larger access opening is likely necessary at salons for performing manicures. The experiments did not consider worker or customer movement and only measured a potential exposure from two direct linear coughs. The OPCs collected data at 6-second intervals, so a travel time associated with the aerosolized particles could not be calculated. The cough simulator did not produce many particles in the larger size ranges. We collected data for 10 minutes after each cough. Since smaller particles remain in the air for longer, the two ACH we used aided in removing the small particles and prevented them from migrating and settling. Finally, masks or other transmission control methods were not considered in this work. Barriers were solely considered to assess this engineering control. Other control methods used in combination with barriers would likely provide additional reductions in exposure.

In summary, laboratory simulations demonstrate that barriers can reduce cough particle exposures in locations where workers interact with others and especially for situations where there is extended interaction time (e.g., nail salons). Barriers can be used in conjunction with other prevention recommendations, e.g., strategies to improve ventilation, vaccination, and masks. While barriers designed to be at least 9 cm (3.5”) above the height of the expected customer’s mouth were shown to be effective, barriers that were 39 cm (15.5”) above the customer’s mouth were more effective. Efficiency above 71% for the smallest particles can be achieved for most workers by using barriers that extend 183 cm (72”) from the floor for standing interactions and 150 cm (59”) from the floor for sitting interactions. The most effective width was 91 cm (36”) for the scenarios studied; wider barriers were not shown to improve barrier efficiency Barriers taller than 39 cm (15.5”) above the customer’s mouth height were not shown to improve barrier efficiency.

## Data Availability

All the data that has been tabulated in the manuscript can be provided in a more accessible format.

## ACKNOWLEDGMENTS

The authors would like to thank William Lindsley, Duane Hammond, Kevin Menchaca, Ken Mead, Brian Curwin and Matt Dahm with their assistance and feedback throughout the experimental process.

## Disclaimer

The findings and conclusions in this report are those of the authors and do not necessarily represent the official position of the National Institute for Occupational Safety and Health (NIOSH), US Centers for Disease Control and Prevention (CDC). Mention of any company or product does not constitute endorsement by NIOSH, CDC.

## Author contributions

JB created the experimental design and performed the experiments. CE assisted in the experimental design and analysis of the data. ICC analyzed the data, wrote the results and generated the tabulated data. DN assisted with the ventilation work and training on the experimental equipment. JB, CE and ICC interpreted the results.

## Supplemental

**Table S1.** Collection efficiency for large sized particles (3.5µm – 6.25µm) in standing and sitting scenarios.

|  | Large Sized Particles (3.5µm – 6.25µm) |  |  |
| --- | --- | --- | --- |
|  | Number of Experiments | Median Efficiency (Range) | P-value |
| **Barrier in Standing Scenario** |  |  |  |
| No Barrier | 6 | Reference |  |
| Height (H) from Cough |  |  |  |
| -16.5 cm (-6.5") H | 18 | -28.58 (-17730 – 75.94) | 0.0912 |
| +14 cm (5.5") H | 18 | 66.50 (-84.24 – 91.00) | <.0001 |
| +44.5 cm (+17.5") H | 18 | 69.83 (-238.1 – 98.82) | <.0001 |
| Width (W) Excluding -16.5 cm (-6.5") H |  |  |  |
| 61 cm (24") W | 12 | 57.89 (-130.0 – 89.96) | <.0001 |
| 91 cm (36") W | 12 | 76.98 (-84.24 – 98.82) | <.0001 |
| 122 cm (48") W | 12 | 65.72 (-238.1 – 92.79) | <.0001 |
| Height (H) * Width (W) |  |  |  |
| -16.5 cm (-6.5") H * 61 cm (24") W | 6 | -22.60 (-139.7 – 75.94) | 0.3816 |
| -16.5 cm (-6.5") H * 91 cm (36") W | 6 | -29.60 (-1603 – 42.47) | 0.2878 |
| -16.5 cm (-6.5") H * 122 cm (48") W | 6 | -69.79 (-17730 – 12.71) | 0.1857 |
| +14 cm (5.5") H * 61 cm (24") W | 6 | 61.74 (36.31 – 85.49) | 0.0001 |
| +14 cm (5.5") H * 91 cm (36") W | 6 | 81.10 (-84.24 – 91.00) | <.0001 |
| +14 cm (5.5") H * 122 cm (48") W | 6 | 60.74 (-25.21 – 77.77) | <.0001 |
| +44.5 cm (+17.5") H * 61 cm (24") W | 6 | 37.00 (-130.0 – 89.96) | 0.3657 |
| +44.5 cm (+17.5") H * 91 cm (36") W | 6 | 76.40 (8.09 – 98.82) | <.0001 |
| +44.5 cm (+17.5") H * 122 cm (48") W | 6 | 71.06 (-238.1 – 92.79) | 0.0006 |
| **Barrier in Sitting Scenario** |  |  |  |
| No Barrier | 6 | Reference |  |
| Height (H) from Cough |  |  |  |
| +9 cm (3.5") H | 18 | 78.57 (-137.4 – 97.03) | <.0001 |
| +39 cm (15.5") H | 18 | 81.15 (7.85 – 98.74) | <.0001 |
| +70 cm (27.5") H | 18 | 70.77 (-17.50 – 94.98) | <.0001 |
| Width (W) |  |  |  |
| 61 cm (24") W | 18 | 63.94 (-116.6 – 94.64) | <.0001 |
| 91 cm (36") W | 18 | 80.27 (21.09 – 98.74) | <.0001 |
| 122 cm (48") W | 18 | 79.97 (-137.4 – 96.63) | <.0001 |
| Height (H) * Width (W) |  |  |  |
| +9 cm (3.5") H * 61 cm (24") W | 6 | 55.22 (-116.6 – 85.25) | 0.0009 |
| +9 cm (3.5") H * 91 cm (36") W | 6 | 86.96 (59.51 – 97.03) | <.0001 |
| +9 cm (3.5") H * 122 cm (48") W | 6 | 80.76 (-137.4 – 96.19) | 0.0001 |
| +39 cm (15.5") H * 61 cm (24") W | 6 | 83.42 (7.85 – 94.64) | <.0001 |
| +39 cm (15.5") H * 91 cm (36") W | 6 | 80.64 (52.19 – 98.74) | <.0001 |
| +39 cm (15.5") H * 122 cm (48") W | 6 | 78.12 (19.54 – 96.63) | <.0001 |
| +70 cm (27.5") H * 61 cm (24") W | 6 | 56.39 (-17.50 – 85.93) | 0.0010 |
| +70 cm (27.5") H * 91 cm (36") W | 6 | 69.42 (21.09 – 94.98) | <.0001 |
| +70 cm (27.5") H * 122 cm (48") W | 6 | 84.77 (2.10 – 94.51) | <.0001 |

**Table S2.**
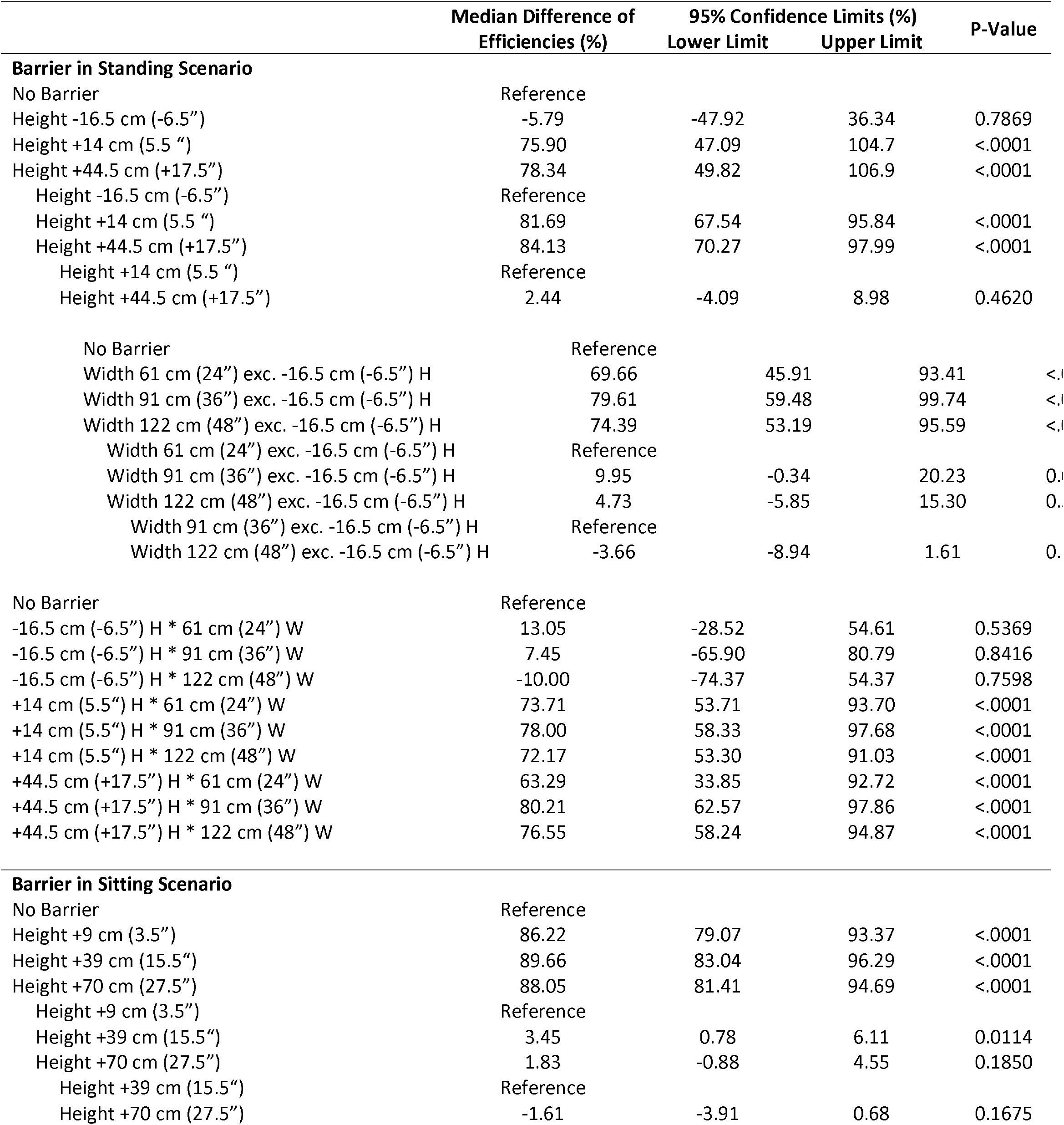
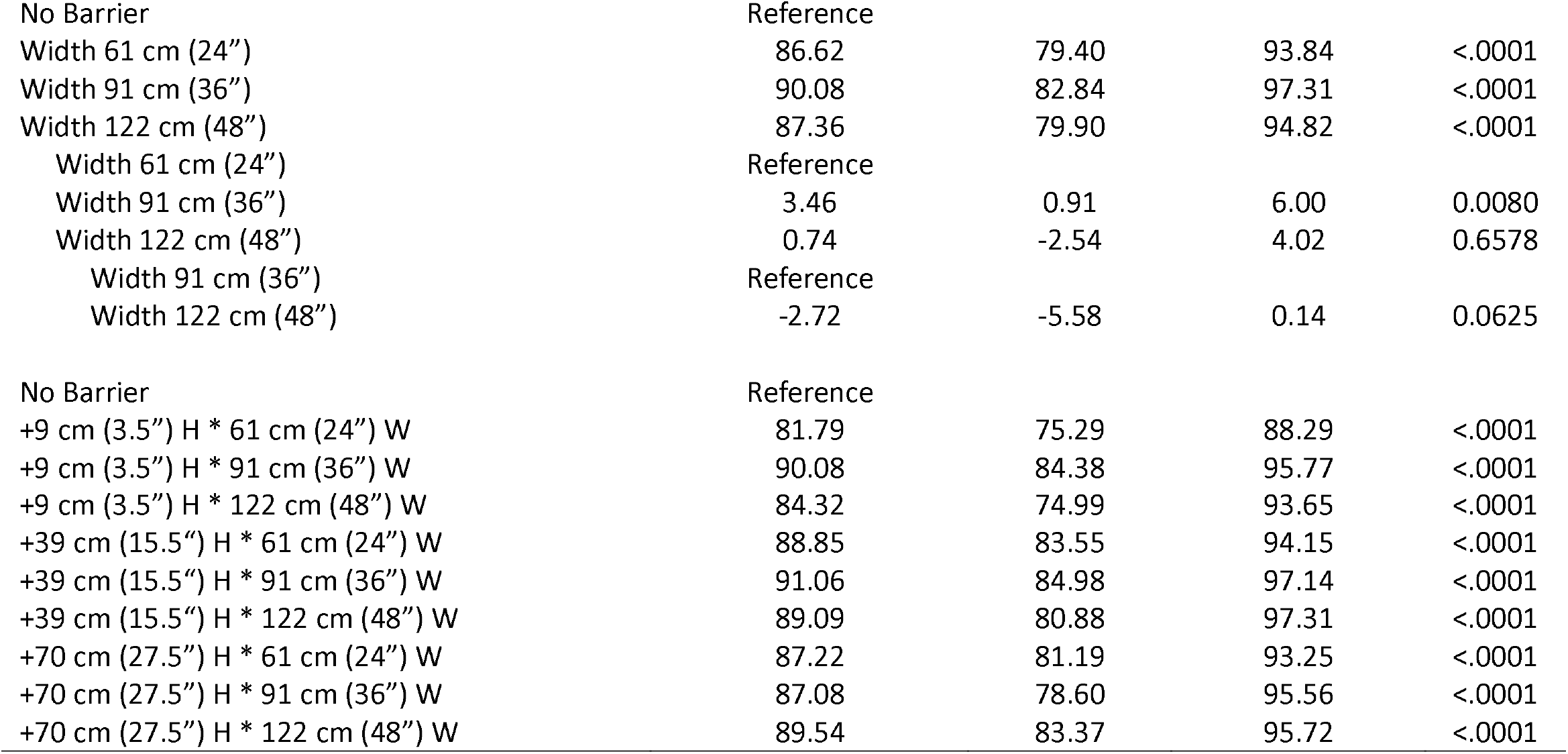
Comparison of barrier settings’ efficiencies for small sized particles (0.35µm – 0.725µm) in standing and sitting scenarios.

**Table S3.** Comparison of barrier settings’ efficiencies for moderate sized particles (0.9µm – 2.5µm) in standing and sitting scenarios.

|  | Median Difference<br>of Efficiencies (%) | 95% Confidence Limits (%) |  | P-Value |
| --- | --- | --- | --- | --- |
|  |  | Lower Limit | Upper Limit |  |
| Barrier in Standing Scenario |  |  |  |  |
| No Barrier | Reference |  |  |  |
| Height -16.5 cm (-6.5") | -36.38 | -100.01 | 27.25 | 0.2612 |
| Height +14 cm (5.5 ") | 65.66 | 27.88 | 103.45 | 0.0007 |
| Height +44.5 cm (+17.5") | 66.67 | 27.95 | 105.40 | 0.0008 |
| Height -16.5 cm (-6.5") | Reference |  |  |  |
| Height +14 cm (5.5") | 102.04 | 79.96 | 124.12 | <.0001 |
| Height +44.5 cm (+17.5") | 103.05 | 80.85 | 125.25 | <.0001 |
| Height +14 cm (5.5") | Reference |  |  |  |
| Height +44.5 cm (+17.5") | 1.01 | -9.07 | 11.09 | 0.8437 |
| No Barrier | Reference |  |  |  |
| Width 61 cm (24") exc. -16.5 cm (-6.5") H | 64.32 | 37.88 | 90.77 | <.0001 |
| Width 91 cm (36") exc. -16.5 cm (-6.5") H | 74.05 | 48.24 | 99.85 | <.0001 |
| Width 122 cm (48") exc. -16.5 cm (-6.5") H | 60.35 | 29.91 | 90.80 | 0.0001 |
| Width 61 cm (24") exc. -16.5 cm (-6.5") H | Reference |  |  |  |
| Width 91 cm (36") exc. -16.5 cm (-6.5") H | 9.72 | 1.40 | 18.04 | 0.0201 |
| Width 122 cm (48") exc. -16.5 cm (-6.5") H | -2.84 | -15.49 | 9.81 | 0.6501 |
| Width 91 cm (36") exc. -16.5 cm (-6.5") H | Reference |  |  |  |
| Width 122 cm (48") exc. -16.5 cm (-6.5") H | -12.56 | -25.00 | -0.13 | 0.0401 |
| No Barrier | Reference |  |  |  |
| -16.5 cm (-6.5") H * 61 cm (24") W | -8.63 | -71.49 | 54.23 | 0.7870 |
| -16.5 cm (-6.5") H * 91 cm (36") W | -15.37 | -129.51 | 98.77 | 0.7910 |
| -16.5 cm (-6.5") H * 122 cm (48") W | -75.02 | -246.44 | 96.41 | 0.3895 |
| +14 cm (5.5") H * 61 cm (24") W | 65.66 | 15.37 | 115.96 | 0.0107 |
| +14 cm (5.5") H * 91 cm (36") W | 72.35 | 14.29 | 130.40 | 0.0148 |
| +14 cm (5.5") H * 122 cm (48") W | 60.35 | -1.17 | 121.88 | 0.0545 |
| +44.5 cm (+17.5") H * 61 cm (24") W | 61.08 | -17.58 | 139.75 | 0.1274 |
| +44.5 cm (+17.5") H * 91 cm (36") W | 76.10 | 24.70 | 127.50 | 0.0039 |
| +44.5 cm (+17.5") H * 122 cm (48") W | 66.31 | 9.49 | 123.13 | 0.0224 |
| Barrier in Sitting Scenario |  |  |  |  |
| No Barrier | Reference |  |  |  |
| Height +9 cm (3.5") | 87.40 | 72.60 | 102.2 | <.0001 |
| Height +39 cm (15.5") | 93.11 | 80.73 | 105.5 | <.0001 |
| Height +70 cm (27.5") | 86.45 | 72.44 | 100.5 | <.0001 |
| Height +9 cm (3.5") | Reference |  |  |  |
| Height +39 cm (15.5") | 5.72 | 0.22 | 11.21 | 0.0416 |
| Height +70 cm (27.5") | -0.09 | -7.54 | 7.36 | 0.9815 |
| Height +39 cm (15.5") | Reference |  |  |  |
| Height +70 cm (27.5") | -5.80 | -11.37 | -0.23 | 0.0414 |
| No Barrier | Reference |  |  |  |
| Width 61 cm (24") | 84.52 | 72.10 | 96.93 | <.0001 |
| Width 91 cm (36") | 92.87 | 82.04 | 103.7 | <.0001 |
| Width 122 cm (48") | 86.76 | 73.16 | 100.4 | <.0001 |
| Width 61 cm (24") | Reference |  |  |  |
| Width 91 cm (36") | 8.35 | 3.38 | 13.32 | 0.0011 |
| Width 122 cm (48") | 2.24 | -6.60 | 11.09 | 0.6181 |
| Width 91 cm (36") | Reference |  |  |  |
| Width 122 cm (48") | -6.11 | -12.05 | -0.17 | 0.0437 |
| No Barrier | Reference |  |  |  |
| +9 cm (3.5") H * 61 cm (24") W | 72.12 | 49.06 | 95.18 | <.0001 |
| +9 cm (3.5") H * 91 cm (36") W | 94.08 | 83.62 | 104.5 | <.0001 |
| +9 cm (3.5") H * 122 cm (48") W | 86.76 | 66.12 | 107.4 | <.0001 |
| +39.4 cm (15.5") H * 61 cm (24") W | 93.11 | 82.07 | 104.1 | <.0001 |
| +39.4 cm (15.5") H * 91 cm (36") W | 92.97 | 82.34 | 103.6 | <.0001 |
| +39.4 cm (15.5") H * 122 cm (48") W | 86.51 | 60.34 | 112.7 | <.0001 |
| +70 cm (27.5") H * 61 cm (24") W | 82.42 | 64.52 | 100.3 | <.0001 |
| +70 cm (27.5") H * 91 cm (36") W | 86.45 | 67.35 | 105.5 | <.0001 |
| +70 cm (27.5") H * 122 cm (48") W | 85.25 | 59.03 | 111.5 | <.0001 |

